# Field performance and cost-effectiveness of a point-of-care triage test for HIV virological failure in Southern Africa

**DOI:** 10.1101/2023.09.18.23295705

**Authors:** Anna Saura-Lázaro, Peter Bock, Erika van den Bogaart, Jessie van Vliet, Laura Granés, Kerry Nel, Vikesh Naidoo, Michelle Scheepers, Yvonne Saunders, Núria Leal, Francesco Ramponi, René Paulussen, Tobias Rinke de Wit, Denise Naniche, Elisa López-Varela

## Abstract

**Introduction:** Antiretroviral therapy (ART) monitoring using viral load (VL) testing is challenging in high-burden, limited-resources settings. Chemokine IP-10 (interferon gamma-induced protein 10) strongly correlates with human immunodeficiency virus (HIV) VL. Its determination could serve to predict virological failure (VF) and to triage patients requiring VL testing. We assessed the field performance of a semi-quantitative IP-10 lateral flow assay (LFA) for VF screening in South Africa, and the cost-effectiveness of its implementation in Mozambique.

**Methods:** A cross-sectional study was conducted between June and December 2021 in three primary health clinics in the Western Cape. Finger prick capillary blood was collected from adults on ART for ≥1 year for direct application onto the IP-10 LFA (index test) and compared with a plasma VL result ≤1 month prior (reference test). We estimated the area under the receiver operating characteristic curves (AUC), sensitivity and specificity, to evaluate IP-10 LFA prediction of VF (VL>1,000 copies/mL). A decision tree model was used to investigate the cost-effectiveness of integrating IP-10 LFA combined with VL testing into the current Mozambican ART monitoring strategy. Averted disability-adjusted life years (DALYs) and HIV infections, and incremental cost-effectiveness ratios were estimated.

**Results:** Among 209 participants (median age 38 years and 84% female), 18% had VF. Median IP-10 LFA values were higher among individuals with VF compared to those without (24.0 vs. 14.6; p<0.001). The IP-10 LFA predicted VF with an AUC=0.76 (95% confidence interval (CI) 0.67–0.85), 91.9% sensitivity (95%CI 78.1%–98.3%) and 35.1% specificity (95%CI 28.0%– 42.7%). Integrating the IP-10 LFA in a setting with 20% VF prevalence and 61% VL testing coverage could save 13.0% of costs and avert 14.9% of DALYs and 55.7% new HIV infections. Furthermore, its introduction was estimated to reduce the total number of routine VL tests required for ART monitoring by up to 68%.

**Conclusions:** The IP-10 LFA is an effective VF triage test for routine ART monitoring. Combining a highly sensitive, low-cost IP-10 LFA-based screening with targeted VL confirmatory testing could result in significant healthcare quality improvements and cost savings in settings with limited access to VL testing.

## Introduction

For people living with human immunodeficiency virus (PLHIV) on antiretroviral therapy (ART), viral load (VL) testing is the gold standard approach to timely monitor treatment effectiveness, identify virological failure (VF) and establish the need to switch to a second-line ART regimen [1]. Prior to the roll-out of dolutegravir in 2019, reported VF rates in Sub-Saharan Africa (SSA) varied from 5% to 25% [2,3]. VF has severe clinical consequences due to the resulting immunosuppression; in addition 80-90% of subjects with detectable viremia may eventually harbor viruses resistant to first-line ART [2–4]. In the absence of adequate ART monitoring tools, PLHIV who have VF can remain on failing regimens with clinical deterioration, onward HIV transmission and gradual loss of ART effectiveness on individual and population levels.

The recent and rapid scale-up of ART coverage, with 28.7 million PLHIV receiving ART in 2021 [5], has increased the demand for VL tests. Access to VL testing is limited in many low- and middle-income countries (LMIC) with high HIV burden, particularly in SSA, due to costs and lack of qualified human resources and infrastructure [6,7]. Data from 2018 showed that in 6 SSA countries assessed the mean proportion of PLHIV on ART receiving at least one VL test was below 75%, and <55% in two of them [8]. Enabling routine access to VL testing to promptly detect VF is critical to reduce HIV burden, morbidity and mortality. In LMIC where widespread use of nucleic acid-based VL tests is not a realistic option, reliance on dried blood spots (DBS), point-of-care (POC) technologies, or other alternative specimen types or technologies is necessary to expand access to VL testing.

IP-10 (interferon gamma-induced protein 10) is a human chemokine that strongly correlates with HIV VL [9]. Its assessment in plasma can serve as a highly sensitive screening tool to identify PLHIV on ART at risk of VF [10,11]. Mondial Diagnostics in The Netherlands developed an IP-10 rapid test for finger prick capillary blood samples and plasma, in collaboration with the Barcelona Institute for Global Health (ISGlobal) and the Amsterdam Institute for Global Health and Development (AIGHD). This semi-quantitative lateral flow assay (LFA) consists of a dried detection reagent deposited in a test tube and a dipstick with a test line to measure IP-10 and a control line to confirm assay validity. We aimed to evaluate the field performance of this first IP-10 LFA prototype as a VF screening test in PLHIV on ART in Cape Town, South Africa. We also assessed the cost-effectiveness of implementing the IP-10 LFA in three different ART monitoring algorithms in a setting with limited access to routine VL testing using Mozambique as an example

## Methods

### Study design, setting and population

A cross-sectional study was conducted between June and December 2021 in three primary health clinics, Kraaifontein Community Health Centre, Ivan Toms Centre for health and Bloekombos Clinic, in the Cape Metro, Western Cape, South Africa. The Cape Metro district has approximately 48,400 PLHIV on ART, a relatively high VL coverage of 72-89% [12,13], and an overall VL suppression rate, defined as undetectable VL, of 84% among individuals with VL result [14]. HIV treatment and care are offered free of charge and VL testing for ART monitoring is recommended to be performed annually [15]. We strategically selected these clinics due to their relatively high programmatic VL coverage, which facilitated the effective evaluation of the IP-10 LFA’s field performance by utilizing existing VL data and infrastructure.

PLHIV aged ≥18 years on ART for at least 12 months without treatment interruptions in the previous 3 months attending the clinic were pre-screened for study participation pursuing an enriched enrolment of PLHIV experiencing VF to ensure 85% statistical power. Assuming a VF prevalence of 25% and a sensitivity of the IP-10 LFA above 90%, we calculated that a sample size of 35 PLHIV experiencing VF would provide a significant level of 0.05 and a confidence interval (CI) with a half-width of 0.1. Individuals presenting with symptoms suggestive of COVID-19, such as cough, shortness of breath, sore throat, anosmia, ageusia, conjunctivitis, and weight loss >1.5kg, were excluded [16].

### Study visit procedures

After obtaining informed consent, a research assistant collected participants’ socio-demographic and clinical information by using an electronic survey. Each participant underwent phlebotomy and finger prick capillary blood testing with the IP-10 LFA (index test) performed by a trained nurse.

The IP-10 LFA was performed in a controlled temperature environment as follows: i) 50 µL of assay fluid were dispensed into the test tube, followed by 2 µL of freshly collected capillary blood using a precision pipette; ii) the dipstick was immersed into the reaction mixture for 20 minutes; iii) after time elapsed, the dipstick was removed from the test tube and added to the clean, portable, battery-operated LFA reader (Cube Reader, Chembio Diagnostics, Germany), which provided a numeric readout in arbitrary units (scale: 0-10,000, 0.1 increments) proportional to the IP-10 concentration in the test sample. The IP-10 LFA result was not shared with the participant.

If the participant had a routine VL completed in the 30 days prior to visit, this result was accessed and captured in the study database. If no routine VL result was available, the study team completed a VL test through using an aliquot of the venous blood collected.

Anti-coagulated venous blood samples were transported to the Clinical Laboratory Services in Cape Town where plasma was separated and cryopreserved within 4 hours of sample collection, prior to being shipped to Mondial Diagnostics for further test validation.

### Laboratory procedures

Plasma HIV RNA levels (VL result) were determined according to routine practice using the Abbott Real-Time HIV1/2 Polymerase Chain Reaction (PCR) (reference test). Quantitative measurement of IP-10 concentrations in plasma samples was conducted at Mondial Diagnostics using the Human CXCL10/IP-10 Quantikine ELISA kit (R&D Systems, USA), according to manufacturer’s specifications. Plasma specimens were also blindly assayed with the IP-10 LFA using 1 µL of plasma sample from the same test batch as the one used for capillary blood at study sites. The outcome was determined by Cube Reader as well as visually by two independent readers (scale: 0-4, 0.5 increments). When the Cube Reader results differed ≥2-fold between plasma and capillary blood, the IP-10 LFA was repeated, and the average result of both tests was included in the analysis. The IP-10 LFA was validated using both capillary blood and plasma against HIV RNA levels and ELISA IP-10 measurements.

### Statistical analysis

Proportions for categorical variables and the median and interquartile range (IQR) for continuous variables were calculated and compared using X2 and nonparametric Mann-Whitney U test, respectively. Spearman test was used to assess correlation coefficients for continuous variables. Logistic regression analysis with penalized likelihood was performed to assess the capacity of the IP-10 LFA and the socio-demographic and clinical variables to predict VF [17], defined as VL>1,000 copies/mL. A multivariable logistic regression model was built by including all variables with a p-value <0.20 in the bivariable analyses (IP-10 LFA values were fixed), followed by backward stepwise selection, where variables with p-values <0.05 could enter the model whereas a p-value <0.10 was required to be retained. A 5-fold cross-validation was used to estimate average accuracy. Receiver operating characteristic analyses were conducted to obtain the area under the curve (AUC), as well as sensitivity, specificity and predictive values for different IP-10 LFA cut-off values. Optimal cut-off values to screen for VF were selected prioritizing a sensitivity above 90%, as the IP-10 LFA was validated as a triage test. Stata version 16 was used for the analyses.

### Cost-effectiveness analysis

We developed a decision tree model (Figure 1) to compare the current standard of care of using VL testing only for ART monitoring (Strategy 1, Supplementary Figure 1) with 3 alternative strategies integrating the IP-10 LFA combined with a confirmatory VL test (Supplementary Figure 2): i) IP-10 LFA + VL test performed immediately after an IP-10 positive result (Strategy 2a), ii) IP-10 LFA + VL test performed 3 months after an IP-10 positive result and after receiving enhanced adherence counselling (EAC) (Strategy 2b), and iii) IP-10 LFA + a second IP-10 LFA 3 months after an IP-10 positive result and EAC + VL test performed only among clients who test positive for the second IP-10 LFA (Strategy 2c). Considering that the IP-10 LFA was developed to be implemented in settings where access to VL testing is limited, we assessed its cost-effectiveness in Mozambique, where the global VL testing coverage was 61% with a VF prevalence of 20% in 2021 [18].

**Figure 1.**
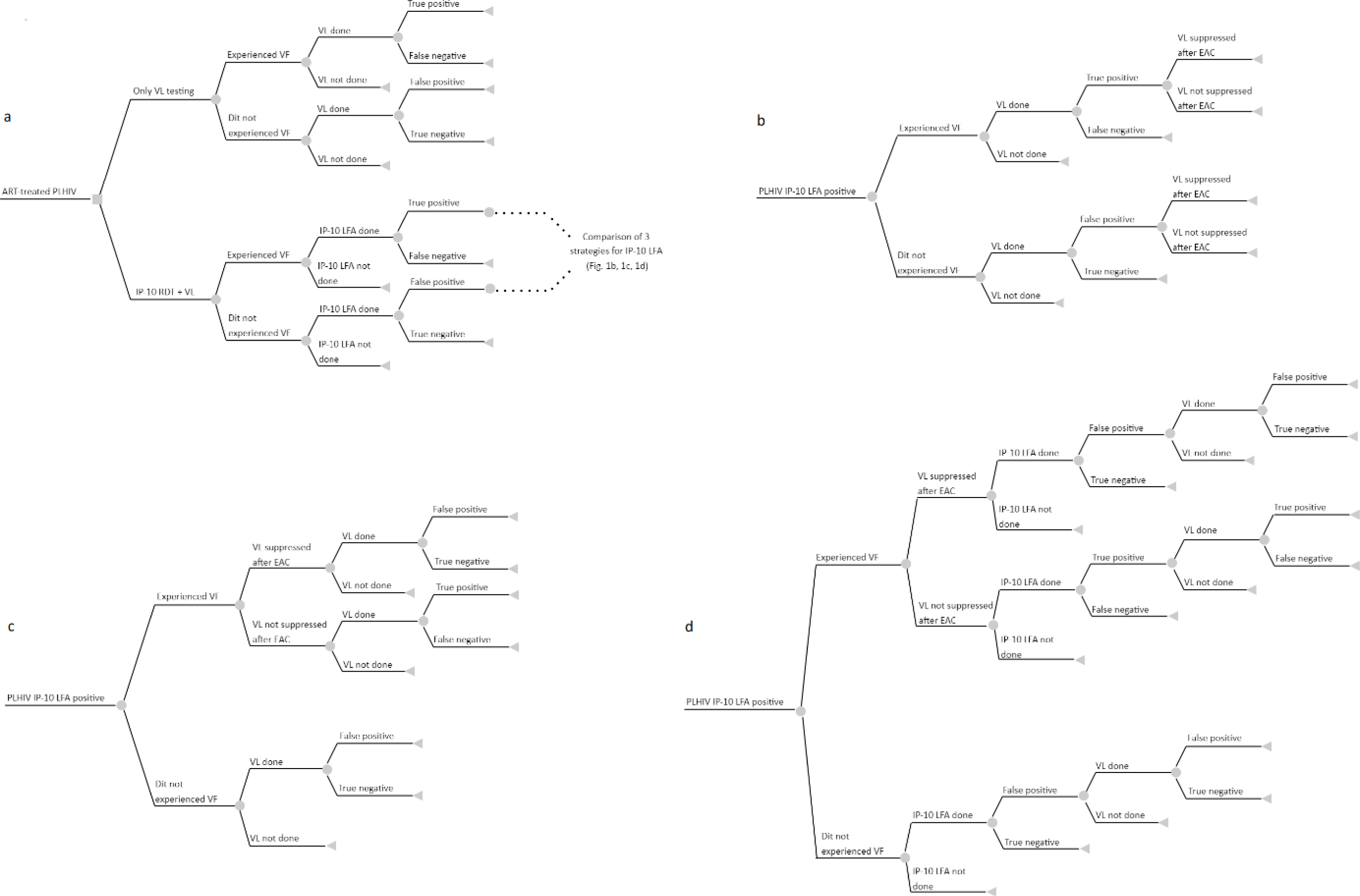
1a) A decision tree model for antiretroviral therapy (ART) monitoring comparing the standard of care of using viral load (VL) testing only in a two-step algorithm recommended by the World Health Organization with three proposed algorithms where the IP-10 lateral flow assay (LFA) is integrated combined with VL confirmatory testing: 1b) immediately after performing the IP-10 LFA; 1c) 3 months later after receiving enhanced adherence counselling (EAC); 1d) 3 months later after receiving EAC and after a second positive IP-10 LFA result. Each branch depicts individual steps of ART monitoring algorithm and is conditional on the previous step. The grey square node represents a decision node, grey circular nodes represent chance nodes, and grey triangles illustrate terminal nodes. PLHIV: people living with HIV, VF: virological failure.

The analysis was conducted from the healthcare system perspective. We adopted a life-time horizon and simulated a cohort of 1,000 adults living with HIV who initiated ART at 25 years of age. Effects on health were measured using disability-adjusted life years (DALYs) averted, and new HIV infections averted (HIA). DALYs were derived from the disability weights according to the Global Burden of Disease Study 2019 [19]. We made the assumption that transmission only occurred from our cohort of 1,000 individuals to a number (n) of other individuals, with n taking values of 1 or 4 (low and high transmission scenarios, respectively). We assumed that all PLHIV with undiagnosed VF, and therefore untreated, generated new HIV cases. Costs included treatment (both first and second-line ART regimens) and monitoring, and we used unit costs of US$3 for IP-10 LFA and of US$51 for VL test (including reagents, transport and staff) [20]. Costs were expressed in 2022 US$ and we applied a 3% discount rate [21,22]. All data used are detailed in Supplementary Table 1. Results were expressed as incremental cost-effectiveness ratios (ICER), calculated as the additional costs to avert one disability-adjusted life year (DALY) or a new HIV infection. A one-way sensitivity analysis was undertaken to investigate the effect of uncertainty in model parameters. Microsoft Excel 2019 was used for the analyses.

### Ethical considerations

The study protocol was approved by the University Health Research Ethics Committee in Cape Town (Approval Nr. N20/02/25) and the Clinical Research Ethical Committee of the Hospital Clinic in Barcelona (Approval Nr. HCB/2021/0656). All study participants completed informed consent.

## Results

### Participant characteristics

We enrolled 209 participants with a median age of 38 years (IQR, 31-44), of whom 83.7% (n=175) were women (Table 1). Median time since HIV diagnosis was 7 years (IQR, 4-10) and median time on ART was 6.5 years (IQR, 4.2-9.4). The most common treatment regimen was Tenofovir/Lamivudine/Dolutegravir (83.3%), and 87.1% reported not missing any ART dose during the last month.

**Table 1.**
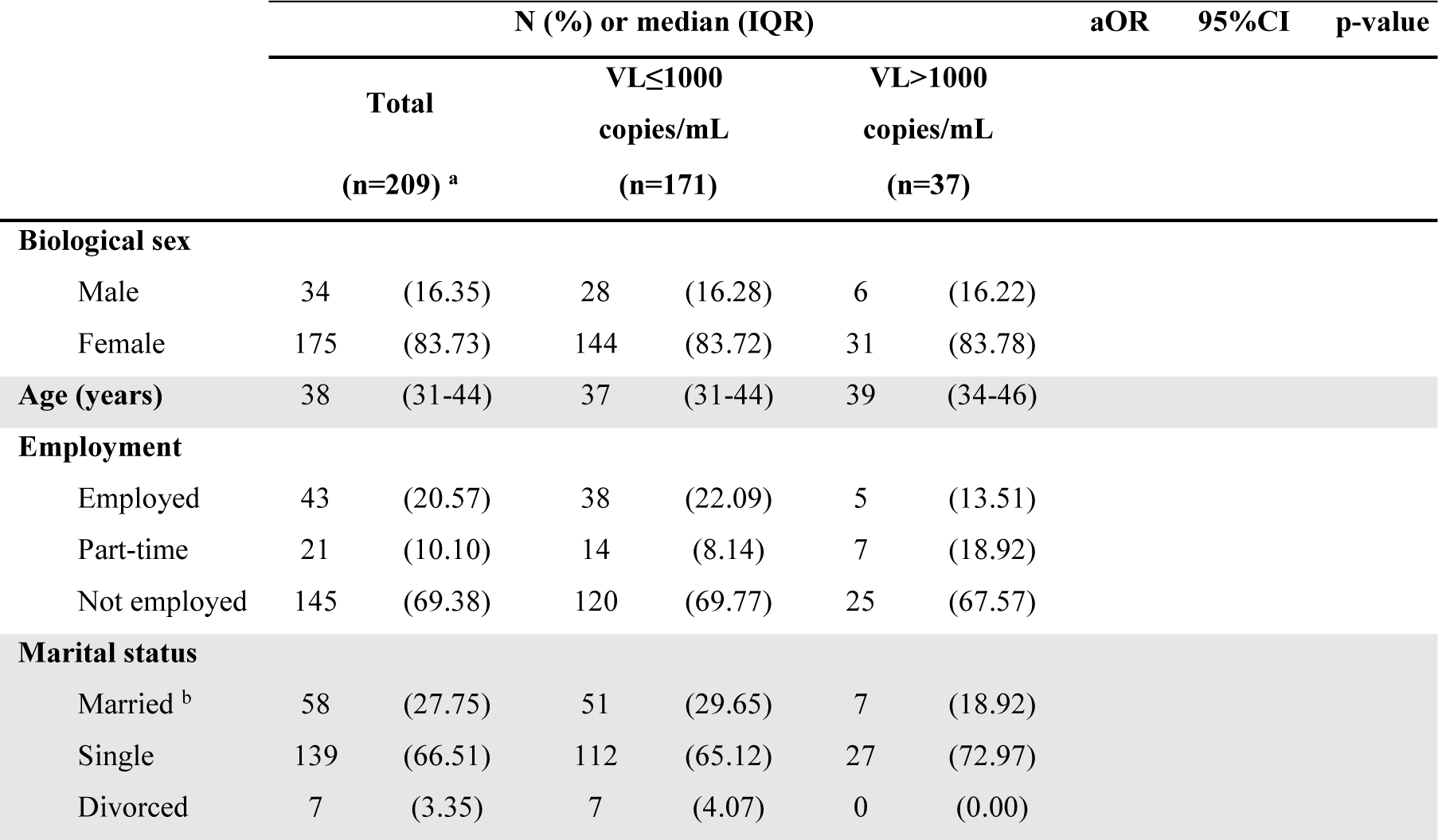
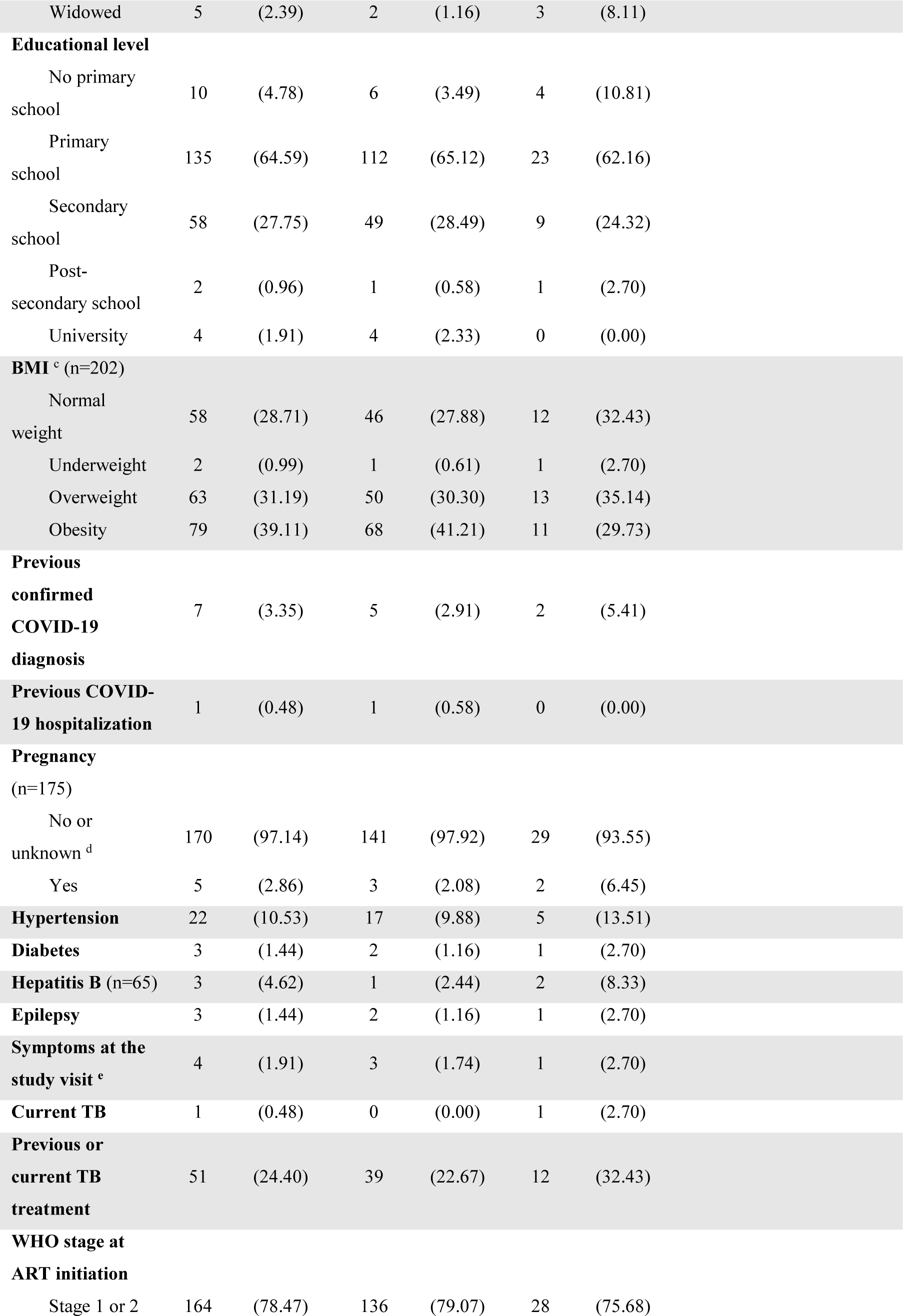
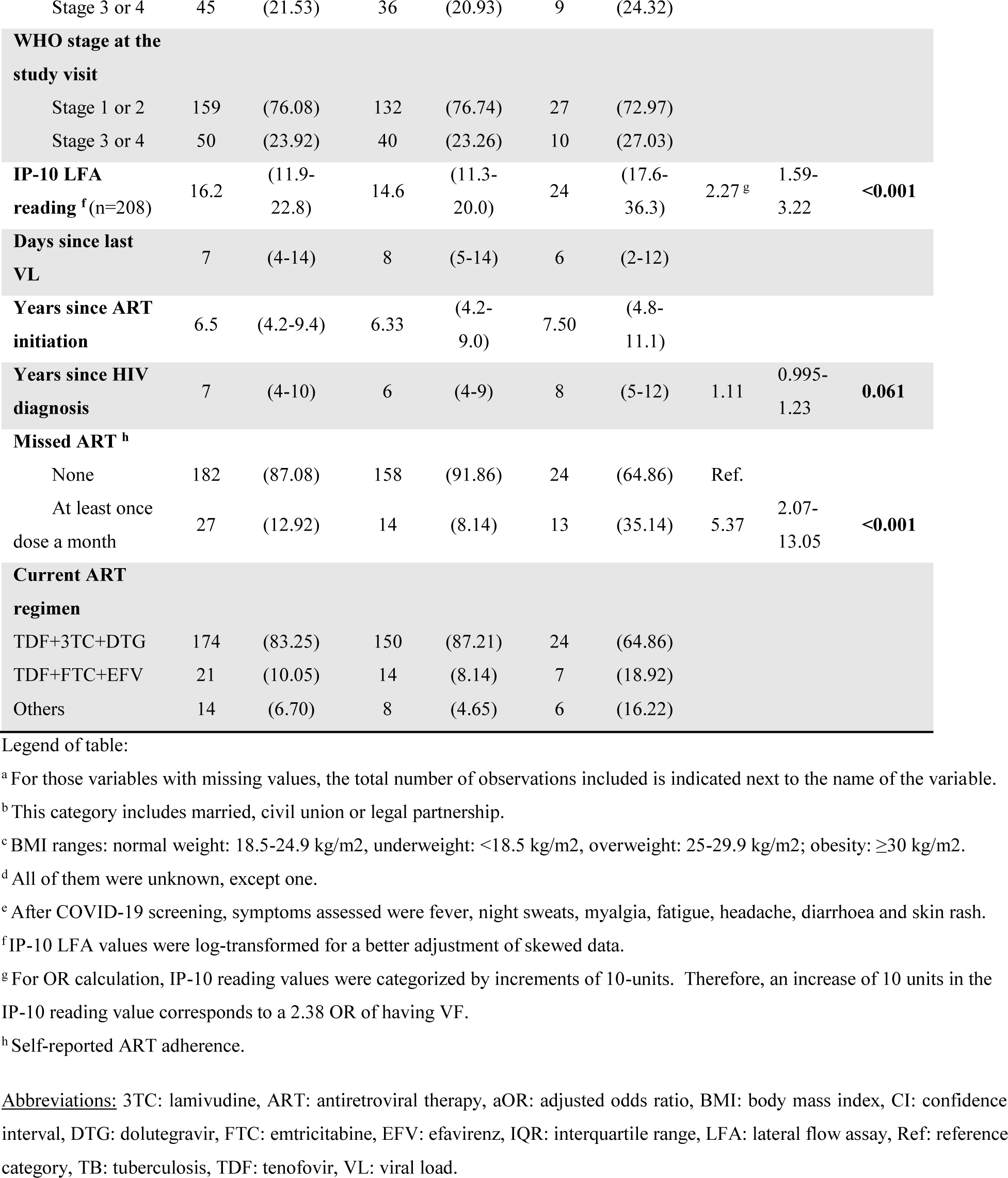
Socio-demographic and clinical characteristics associated with virological failure (VF) (viral load >1,000 copies/mL) among study participants (n=209). Adjusted odds ratios from logistic regression analyses.

Median time since last VL determination was 7 days (IQR, 4-14). A total of 17.7% (95% CI, 12.5-22.9) had VF and among those, median VL was 10,615 copies/mL (IQR, 3,698-49,311). We excluded one participant from the evaluation of the IP-10 LFA performance analysis due to an invalid reading value.

### Field performance

Median IP-10 LFA reading values, hereinafter referred to as ‘IP-10 LFA values’, were significantly higher among individuals with VF (24.0 vs 14.6; p<0.001). Among individuals (n=57) with detectable VL (>50 copies/mL), IP-10 LFA values showed a significant moderate correlation with VL (ρ=0.46, p<0.001).

Multivariable analysis revealed that IP-10 LFA values, self-reported ART adherence and time since HIV diagnosis were associated with VF (Table 1). The univariable analysis (Supplementary Table 2) showed that IP-10 LFA values were associated with VF (odds ratio (OR), 2.38 per 10 units-increase; 95%CI, 1.66-3.40). The estimated AUC was 0.76 (95%CI, 0.67-0.85) and the averaged 5-fold cross validated AUC was 0.77 (95%CI, 0.64-0.84).

Since no differences were found between the AUC of univariable and multivariable models, the former was selected for assessing the accuracy of IP-10 LFA in predicting VF at various cut-off values (Figure 2a). Using an IP-10 LFA value of ≥12.8, the model identified VF with 91.9% sensitivity (95%CI, 78.1%-98.3%) and 35.1% specificity (95%CI, 28.0%-42.7%) (Figure 2b) resulting in a positive predictive value (PPV) of 23.4% (95%CI, 16.8%-31.2%) and a negative predictive value (NPV) of 95.2% (95%CI, 86.7%-99.0%) in the current study population (17.7% VF prevalence) (Supplementary Figure 3).

**Figure 2.**
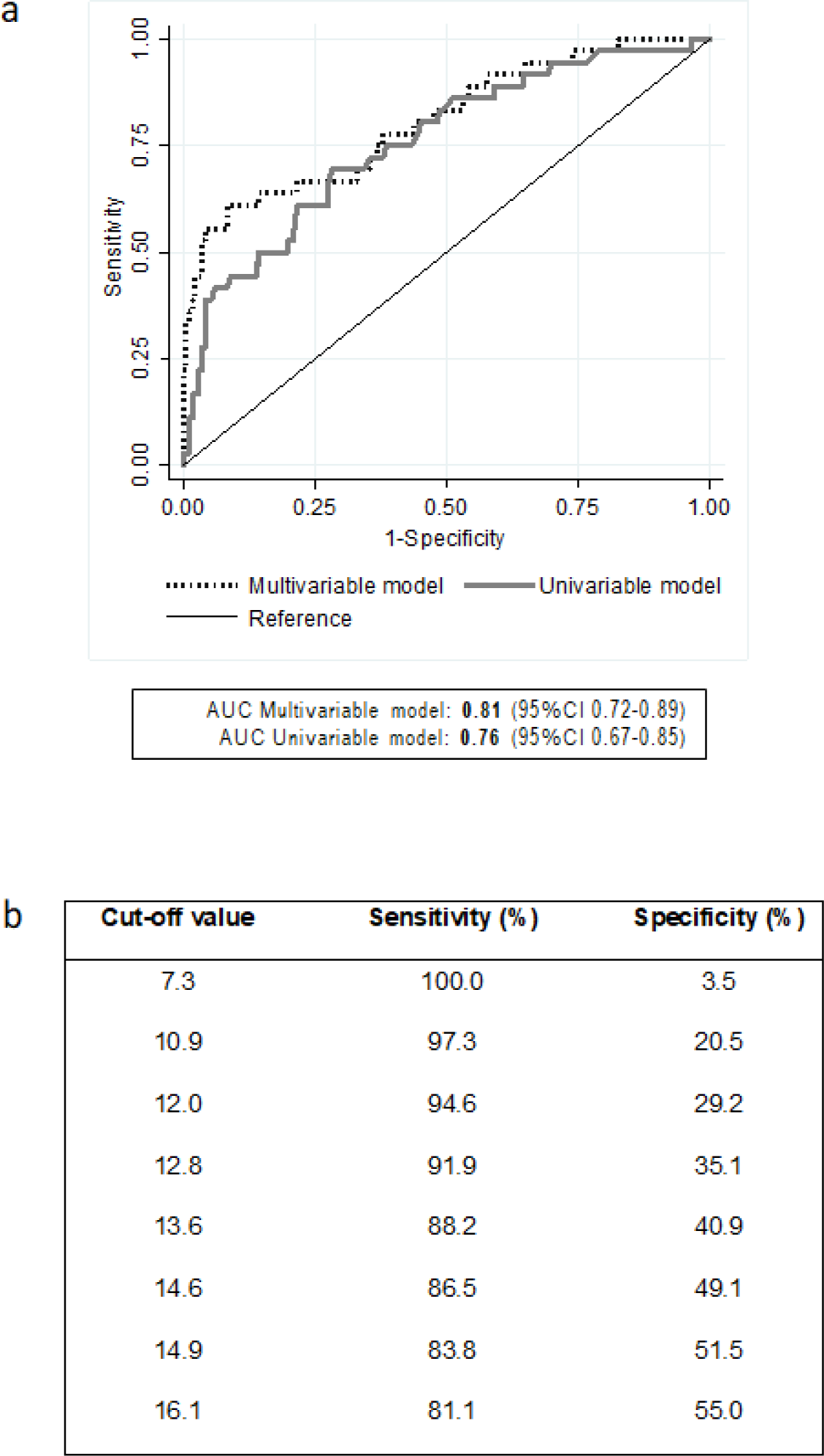
Performance of the IP-10 lateral flow assay (LFA) reading values (arbitrary units, Cube Reader) models in predicting virological failure (VF). (a) Comparison between the area under the curve (AUC) for univariable and multivariable models. (b) Univariable model cut-off IP-10 LFA reading values with their respective sensitivity and specificity values.

### Laboratory validations

Validation of the IP-10 LFA either on capillary blood or plasma against IP-10 ELISA demonstrated a good level of correlation between the two assays, which was higher when the IP-10 LFA was performed on plasma (ρ=0.89, p<0.001) compared to finger prick capillary blood (ρ=0.74, p<0.001) (Supplementary Figure 4). Overall, IP-10 LFA values measured in plasma were lower than in capillary blood but remained significantly higher among individuals with VF (median values of 16.6 vs. 11.9; p<0.001).

The AUC of the IP-10 LFA in plasma to detect VF was similar to capillary blood, regardless of whether the test was read by Cube Reader (0.77; 95%CI, 0.67-0.86) or visually (0.78; 95%CI, 0.69-0.87). When using a cut-off value of 10.7 for the Cube Reader and of 1.5 for visual results, the IP-10 LFA on plasma predicted VF with the same sensitivity as in blood, but the specificity increased to 43.6% (95%CI, 36.1%-51.4%) and 48.8% (95%CI, 41.2%-56.6%), respectively (Table 2).

**Table 2.** Diagnostic accuracy of the IP-10 LFA compared to Real Time-Polymerase Chain Reaction (PCR) for the detection of virological failure (VF), defined as viral load >1,000 copies/mL.

|  | Se<br>(95% CI) | Sp<br>(95% CI) | PPV<br>(95% CI) | NPV<br>(95% CI) | Kappa values<br>(95% CI) |
| --- | --- | --- | --- | --- | --- |
| IP-10 LFA (capillary blood, Cube Reader) | 91.9<br>(78.1 – 98.3) | 35.1<br>(28.0 – 42.7) | 23.4<br>(16.8 – 31.2) | 95.2<br>(86.7 – 99.0) | 0.126<br>(-0.081 – 0.333) |
| IP-10 LFA (plasma, Cube Reader) | 91.9<br>(78.1 – 98.3) | 43.6<br>(36.1 – 51.4) | 26.0<br>(18.7 – 34.3) | 96.2<br>(89.2 – 99.2) | 0.178<br>(-0.011 – 0.366) |
| IP-10 LFA (plasma, visual score) | 91.9<br>(78.1 – 98.3) | 48.8<br>(41.2 – 56.6) | 27.9<br>(20.1 – 36.7) | 96.6<br>(90.3 – 99.3) | 0.214<br>(0.037 – 0.392) |

Legend of table:
IP-10 LFA values were log-transformed for a better adjustment of skewed data.
Abbreviations: CI: confidence interval, LFA: lateral flow assay, NPV: negative predictive value, PPV: positive predictive value, Se: Sensitivity, Sp: Specificity, VF: virological failure, VL: viral load.

### Cost-effectiveness analysis

Costs and health outcomes associated with various ART monitoring strategies are reported in Table 3, ranked by costs in ascending order. In a low transmission scenario (1:1), Strategy 2b was the least expensive strategy with the greatest number of DALYs averted. Compared to the current standard of care (Strategy 1), Strategy 2b could reduce 1.1% of life-time costs and 10.1% of the DALYs associated with HIV. In a high transmission scenario (1:4), Strategy 2b was still the least expensive, associated not only with 13.0% lower costs compared to current standard of care, but also with 14.9% DALYs and 55.7% new HIV infections fewer. Compared to Strategy 2b, Strategy 2c was more expensive, but offered better health outcomes, so it was associated to an ICER of US$1,995and US$2,827 per DALY and HIA averted, respectively. Strategies 2b and 2c were estimated to reduce up to 46.9% and 67.9% respectively, of the total routine VL tests necessary for ART monitoring.

**Table 3.** Health (DALYs and new HIV infections averted) and economic outcomes of antiretroviral therapy (ART) monitoring strategies. Strategies are listed and compared in order of lowest to highest cost, by low and high HIV transmission scenarios (1:1 and 1:4, respectively).

| Strategy | Total costs<br>(2022 US\$) | Difference<br>in costs, US\$ | Total<br>DALYs | DALYs<br>averted | ICER<br>(US\$/DALY<br>averted) | Total new<br>HIV<br>infections | New HIV<br>infections averted | ICER (US\$/HIV<br>infection averted) |
| --- | --- | --- | --- | --- | --- | --- | --- | --- |
| **Low transmission scenario (1:1)** |  |  |  |  |  |  |  |  |
| IP-10 LFA + VL test 3 months later<br>after an IP-10 positive result and EAC<br>(Strategy 2b) | 3,897,852 | - | 5,886.1 | - | - | 40.7 | - | - |
| IP-10 LFA + VL test immediately after<br>an IP-10 positive result (Strategy 2a) | 3,912,788 | 14,936 | 5,998.9 | -112.8 | Dominated by<br>Strategy 2b | 43.4 | -2.7 | Dominated by<br>Strategy 2b |
| Using VL testing only (Strategy 1) | 3,940,416 | 42,546 | 6,550.7 | -664.6 | Dominated by<br>Strategy 2b | 91.8 | -51.1 | Dominated by<br>Strategy 2b |
| IP-10 LFA + VL test 3 months later<br>after an IP-10 positive result and EAC,<br>and after a second positive IP-10 LFA<br>(Strategy 2c) | 4,032,908 | 135,056 | 5,901.9 | -15.9 | Dominated by<br>Strategy 2b | 34.9 | 5.8* | 24,607* |
| **High transmission scenario (1:4)** |  |  |  |  |  |  |  |  |
| IP-10 LFA + VL test 3 months later<br>after an IP-10 positive result and EAC<br>(Strategy 2b) | 4,384,756 | - | 6,227.9 | - | - | 162.8 | - | - |
| IP-10 LFA + VL test immediately after an IP-10 positive result (**Strategy 2a**) | 4,431,989 | 47,233 | 6,363.4 | -135.5 | Dominated by Strategy 2b | 173.6 | -10.8 | Dominated by Strategy 2b |
| IP-10 LFA + VL test 3 months later after an IP-10 positive result and EAC, and after a second positive IP-10 LFA (**Strategy 2c**) | 4,450,373 | 65,517<sup>†</sup> | 6,195.0 | 32.9<sup>†</sup> | 1,995<sup>†</sup> | 139.5 | 23.2<sup>†</sup> | 2,827<sup>†</sup> |
| Using VL testing only (**Strategy 1**) | 5,038,771 | 654,015 | 7,321.8 | -1,093.9 | Dominated by Strategy 2b | 367.1 | -204.4 | Dominated by Strategy 2b |

Legend of table:
A dominated scenario occurs when the comparator is less costly but more effective. Abbreviations: DALY: disability-adjusted live years, EAC: enhanced adherence counselling,
ICER: incremental cost-effectiveness ratio, LFA: lateral flow assay, VL: viral load. <sup>†</sup>Estimated by comparing Strategy 2c against Strategy 2b, after eliminating dominated strategies.

When comparing Strategy 2c against 2b, the one-way sensitivity analysis (Figure 3) showed that in a high transmission scenario when using the minimum value estimated for VF (10%) in Mozambique in 2021 [13], Strategy 2c was even more expensive but more effective than 2b with an ICER over US$7,500/DALY averted. However, when using the maximum value estimated for VF prevalence of 31% [13], Strategy 2c dominated Strategy 2b as it had lower costs and improved outcomes. Strategy 2c also dominated when decreasing VL coverage to 48% or VL specificity to 96.1%, and when increasing IP-10 LFA sensitivity to 98.3%. Lastly, we found that as the VL suppression rate after EAC decreased, the ICER comparing Strategy 2c against 2b also decreased. Results are detailed in Supplementary Table 3.

**Figure 3.**
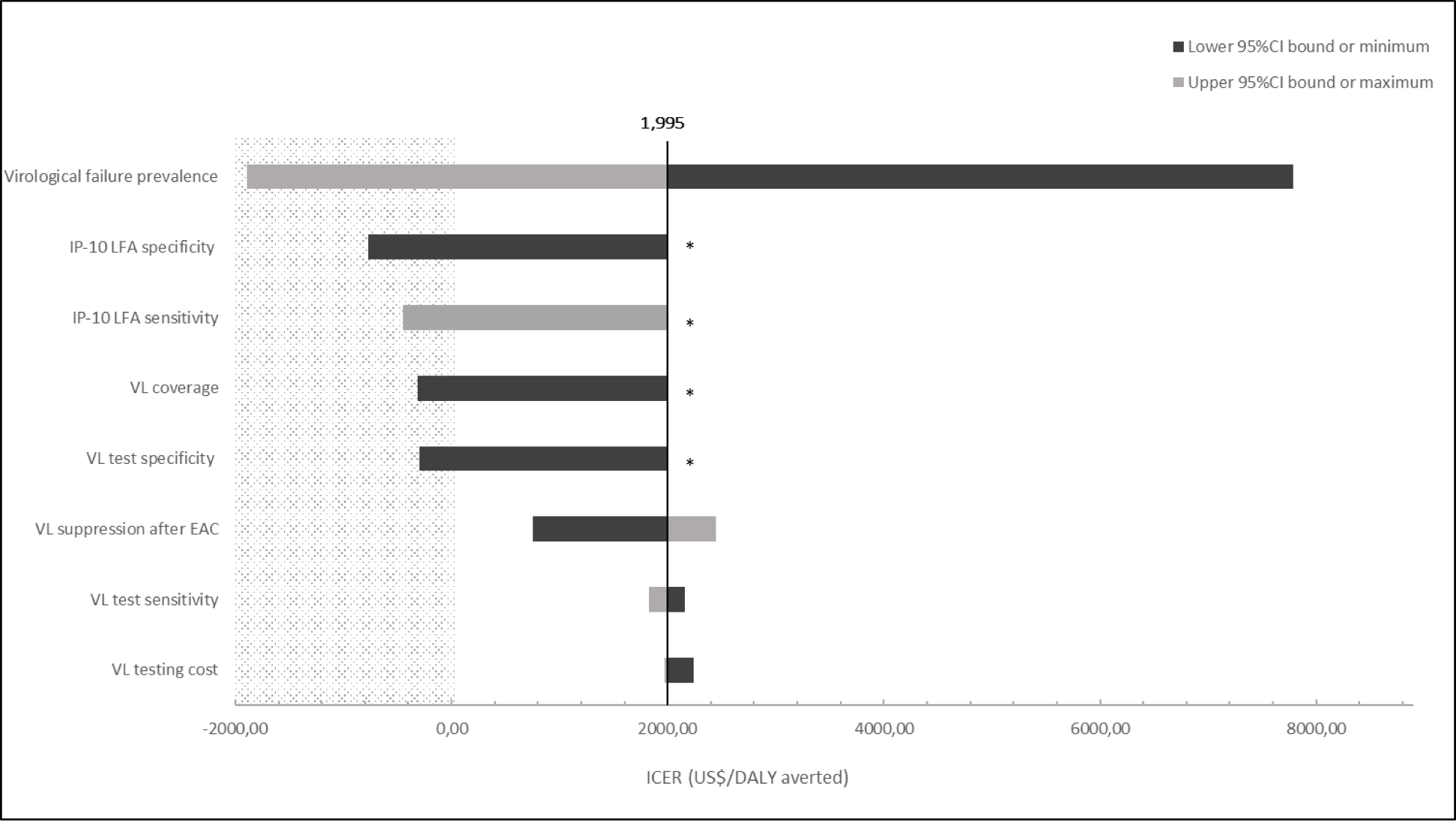
Tornado diagram illustrating fluctuations on the incremental cost-effectiveness ratios (ICER) between the minimum (low) and maximum (high) values of individual model parameters in univariate sensitivity analysis in a high transmission scenario (1:4). ICERs were estimated comparing Strategy 2c against Strategy 2b. Value for each parameter is substituted one by one. Dark grey colour represents the low value (lower 95% CI bound or minimum), while light grey colour represents the high value (upper bound of the 95% CI or maximum) of each of the parameters. The black vertical solid line represents the baseline value of the ICER (1,995 US$/DALY averted). The area shaded dotted grey represents a dominant scenario by Strategy 2c (lower costs and improved outcomes). Parameters that have the highest impact on the model are shown at the top, while the least impact is displayed at the bottom. *Strategy 2b dominated Strategy 2c (lower costs and improved outcomes), so the ICER was not estimated. DALY: disability-adjusted live years, EAC: enhanced adherence counselling, LFA: lateral flow assay, VL: viral load.

## Discussion

This study demonstrated that the IP-10 LFA performed as a highly sensitive VF screening test and a cost-saving tool for ART monitoring in settings with limited access to VL testing. Integrating the IP-10 LFA combined with a confirmatory VL test would allow to rule out VF in PLHIV who are stable and test negative for IP-10 LFA, thus allowing to prioritize available VL capacity for those for whom treatment failure is suspected. Compared to the standard of care, the implementation of IP-10 LFA would expand access to better health outcomes for lower costs. The choice of using IP-10 LFA once (Strategy 2b) or twice (Strategy 2c) in the WHO-aligned algorithm is context-specific and would depend on local factors as well as the cost-effectiveness threshold.

Even though most of PLHIV with VF were identified by IP-10 testing, up to 8% yielded a false negative result. Especially in LMIC the consequences of such VF misdiagnosis need to be weighed against the morbidity and mortality risks associated with a higher proportion of PLHIV not having access to ART monitoring at all. Self-reported treatment adherence was a strong predictor of VF in this study; further investigation should explore if its assessment integrated in the algorithm can help identify IP-10 false negative results.

Regarding specificity, IP-10 is not specific to HIV and increases in other infections such as tuberculosis or COVID-19 [23,24]. With a moderate specificity, such as that observed in this study, approximately 43% of participants without VF would be correctly identified as such with the IP-10 test when using plasma, while the remaining 57% without VF would have a positive IP-10 LFA. HIV VL is also sensitive to conditions which activate the immune system such as co-infections and vaccines [25,26]. Nevertheless, based on the proposed implementation algorithms, individuals with a positive IP-10 result would receive EAC followed by a 3-month confirmatory VL testing, a non-invasive intervention of which the only consequence could be to reinforce their adherence to treatment.

This IP-10 LFA prototype performed as well as an ELISA test with a sensitivity >90% and a moderate specificity, which improved when using plasma (43%) instead of capillary blood (35%). This is likely due to the interindividual variation in hematocrit levels that affects the actual plasma volume being tested from a drop of capillary blood. Nevertheless, moderate specificity is a common feature of screening tests which typically require confirmation with a more specific diagnostic test [27].

Based on the economic evaluation, we estimated that the ART monitoring algorithm integrating the IP-10 LFA is superior to the standard of care DBS-based VL testing only in terms of costs and health outcomes. Despite the necessity of confirming positive IP-10 LFA results with VL testing, the introduction of the IP-10 LFA as a VF screening test could reduce more than a half the number of VL tests for routine ART monitoring.

The IP-10 LFA integrated in Strategy 2b was estimated to yield the lowest costs and to avert the greatest number of DALYs in a setting with low HIV transmission. In a high HIV transmission setting, Strategy 2c was demonstrated to be more effective than Strategy 2b, but at the expense of additional costs, US$1,995 for each additional DALY averted and US$2,827for each additional new HIA. Both ICERs were above the recommended cost-effectiveness threshold (CET) of US$250 to US$750 (0.5 to 1.5 times the gross domestic product per capita of Mozambique in 2021 [28,29], and alternative CET estimates based on opportunity costs [30,31]). Strategy 2c was hence not cost-effective when compared to Strategy 2b, although it was still less expensive with better health outcomes than standard of care. However, Strategy 2c demonstrated superiority to 2b (cheaper and more effective) when it was modelled in a scenario with a higher VF prevalence (parameter with the largest impact on the ICER), higher sensitivity of the IP-10 LFA (screening test) and lower specificity and coverage of the VL test (confirmatory test), the key determinants of the utility of a screening program prompting the use of a second screening with the IP-10 LFA [32]. Additionally, Strategy 2c appeared to be potentially cost-effective with an ICER of US$755 when the rate of VL suppression after receiving EAC was lower. This suggests that the use of the IP-10 LFA twice may be cost-effective in settings with a high prevalence of HIV drug resistance.

The major limitation of this study is the small sample size of PLHIV experiencing VF, resulting in relatively wide confidence intervals, especially for the sensitivity and NPV. Another potential limitation may be the interference of unidentified COVID-19 infections, participants with asymptomatic infections may have entered the study, and other undiagnosed co-infections. Future studies should explore the role of co-infections recruiting a higher number of PLHIV with VF to decrease the uncertainty of the estimates. Regarding the cost-effectiveness analysis, the model was static so the number of HIA may be underestimated [33]. Lastly, we found major pricing discrepancies of VL testing across the literature [34–37], but the cost of VL testing had only a minor effect on the ICER.

The IP-10 LFA is a low-cost and easy-to-read rapid test which can be performed by minimally trained personnel. It also offers the advantage of providing results to patients on the same day the test is performed, which can potentially have a positive influence on their behavior and adherence. Therefore, the IP-10 LFA represents a true POC screening test for VF [38] compared with the several POC VL testing technologies developed in recent years, including the WHO-prequalified Cepheid GeneXpert® and Abbott m-PIMA™, which are device-based semi-POC tests with limited portability and increased costs (US$25-US$30, reagents only) [37,39–41]. Moreover, the test read visually performed as well as when using a Cube Reader, with the potential of simplifying its use without the need of an extra device.

The advent of POC technologies coincides with the rollout of the differentiated HIV care models in many LMIC, new less resource-demanding and more client-centered services which typically focused on providing ART to stable PLHIV. The implementation of the IP-10 LFA would expand and improve the access to differentiated HIV care by a quicker triage of PLHIV on ART into differentiated care pathway, same-day results, fewer clinical visits and faster clinical making-decision [42]. The IP-10 LFA may also be amenable to community-based and self-testing, which helps improve access to ART monitoring particularly in hard-to-reach populations, such as people who inject drugs, migrants and PLHIV living in remote or conflict settings [43,44].

## Conclusions

Combining a highly sensitive, low-cost IP-10 LFA-based VF screening with VL confirmatory testing in an optimized algorithm could provide a greatly needed ART monitoring tool with the potential to fill the gap in VL testing access in LMIC. Successful integration of the IP-10 LFA into HIV services must consider PLHIV needs, co-infections, clinic flows, staff training, quality control and supply chain management at decentralized sites. Thus, future studies to evaluate sustainability, acceptability and feasibility of this new IP-10 LFA, as well as to explore its utility in other potential high-risk groups, like children, adolescents, and pregnant women are highly recommended.

## Authors’ contributions

P.B., T.R.W., D.N. and E.L.V. were responsible for conceptualization and study design. K.N., V.N., M.S. and Y.S. recruited subjects and collected and validated clinical data. K.N., V.N., M.S. and Y.S. coordinated sample collection and processing at the field. E.B. and J.V. performed biomarker quantification at the laboratory and validation of the data. A.S.L., E.B. and L.G. performed the statistical analysis. A.S.L., N.L. and F.R. performed the cost-effectiveness analysis. A.S.L., P.B., E.B., L.G., N.L., F.R., D.N. and E.L.V. interpreted the data. A.S.L. drafted the paper. P.B., E.B., M.S., R.P., T.B.W., D.N. and E.L.V, performed the critical data review and revision of manuscript writing. All authors read and approved the final version of the manuscript.

## Acknowledgements

We acknowledge support from the Spanish Ministry of Science and Innovation and State Research Agency through the “Centro de Excelencia Severo Ochoa 2019-2023” Program (CEX2018-000806-S), and support from the Generalitat de Catalunya through the CERCA Program. The authors gratefully acknowledge the staff at the Kraaifontein Community Health Centre, the Ivan Toms Centre for health and the Bloekombos Clinic, in the Cape Metro, South Africa, who worked to collect and manage the data, our research team, collaborators, and especially all communities and participants involved.

## Data Availability Statement

The data that support the findings of this study are available from the corresponding author upon reasonable request.

## Supporting Information

Supporting Information file 1: Supplementary Table 1

Supporting Information file 2: Supplementary Table 2

Supporting Information file 3: Supplementary Table 3

Supporting Information file 4: Supplementary Figure 1

Supporting Information file 5: Supplementary Figure 2

Supporting Information file 6: Supplementary Figure 3

Supporting Information file 7: Supplementary Figures Legends

## List of abbreviations

AIGHD: Amsterdam Institute for Global Health and Development
ART: antiretroviral therapy
AUC: area under the curve
CI: confidence interval
DALY: disability-adjusted live years
DBS: dried blood spots
EAC: enhanced adherence counselling
HIA: HIV infections averted
ICER: incremental cost-effectiveness ratios
IQR: interquartile range
IP-10: interferon gamma-induced protein 10
ISGlobal: Barcelona Institute for Global Health
LFA: lateral flow assay
LMIC: low- and middle-income countries
OR: odds ratio
PCR: Polymerase Chain Reaction
PLHIV: people living with human immunodeficiency virus
POC: point-of-care
PPV: positive predictive value
NPV: negative predictive value
SSA: Sub-Saharan Africa
VF: virological failure
VL: viral load

