## Supplementary Table 1 for "Field performance and cost-effectiveness of a point-of-care triage test for HIV virological failure in Southern Africa"

**Supplementary Table 1. Key model parameters (HIV-related estimates and costs) and sources of data from Mozambique.**

| | **Value or ranged explored** | **Source(s)** |
|---|---|---|
| **HIV-related parameters** | | |
| After ART initiation, probability of experiencing VF (%) | 20.0 (10.0-31.0) [a] | Mozambique Annual Report on HIV-AIDS Related Activities (2021) |
| After a positive IP-10 LFA, probability of experiencing VF (%) | 28.1 | Estimated |
| Life expectancy of PLHIV with suppressed VL (years) | 60.9 | World Bank Open Data (2022) |
| Life expectancy of PLHIV experiencing VF who are diagnosed and treated (years) | 49.0 | McCluskey et al (2017) |
| Life expectancy of PLHIV experiencing VF who are not diagnosed and not treated (years) | 38.0 | Poorolajal et al (2016) |
| After ART initiation, mean time to experiencing VF (years) | 11.0 | McCluskey et al (2017) |
| Disability weight of PLHIV receiving ART [b] | 0.078 | Global Burden of Disease Study (2019) |
| Disability weight of people living with HIV/AIDS, receiving ART [b, c] | 0.274 | Global Burden of Disease Study (2019) |
| Disability weight of people living with HIV/AIDS, not receiving ART [b, d] | 0.582 | Global Burden of Disease Study (2019) |
| **HIV care-related parameters** | | |
| Probability of having a VL test (%) | 61.0 (48.0-77.0) [a] | Mozambique Annual Report on HIV-AIDS Related Activities (2021) |
| Probability of having a VL test after a first screening with IP-10 LFA and receiving EAC (%) | 51.0 | Estimated |
| Probability of having a VL test after a first screening with IP-10 LFA, receiving EAC and having again a second screening with IP-10 LFA (%) | 82.4 | Estimated |
| Probability of having an IP-10 LFA (%) | 100.0 | Assumption |
| VL testing sensitivity (%) [e] | 88.7 (81.1-94.4) [f] | Fajardo et al (2014), Swannet et al (2017) |
| VL testing specificity (%) [e] | 97.8 (96.1-98.8) [f] | Fajardo et al (2014), Swannet et al (2017) |
| IP-10 LFA sensitivity (%) [g] | 91.9 (78.1-98.3) [f] | Estimated |
| IP-10 LFA specificity (%) [g] | 43.6 (36.1-51.4) [f] | Estimated |
| VL suppression after EAC (%) | 45.0 (31.3-66.4) | Diress et al (2019), Bvochora et al (2019) |

| Costs parameters (2022 US$) | | |
|---|---|---|
| VL testing [h] | 51.9 (24.8-54.1) | Korenromp et al (2015) |
| IP-10 LFA | 3 | Estimated |
| EAC (3-monthly sessions) | 11.8 | Data obtained from a HCW |
| Cost per person/year of a PLHIV with suppressed VL (First-line ART drugs) [i] | 311.6 | Korenromp et al (2015) |
| Cost per person/year of a PLHIV with suppressed VL (Second-line ART drugs) [i,j] | 589.9 | Korenromp et al (2015) |
| Cost of a new HIV infection in the first year [k] | 316.3 | Korenromp et al (2015) |
| Cost per person/year of a PLHIV experiencing VF [l] | 543.0 | Pastor et al (2017) |

Legend of table:

[a] Minimum and maximum.

[b] DALYs were derived from the disability weights according to the Global Burden of Disease Study 2019.

[c] Proxy for the disability weight of PLHIV experiencing VF who are diagnosed and treated.

[d] Proxy for the disability weight of PLHIV experiencing VF who are not diagnosed and are not treated.

[e] Measurement of VL using finger prick capillary blood samples and the NucliSENS Easy-Q HIV-1 v2.0 assay. The sensitivity and the specificity were estimated comparing the performance of using dried blood spots from finger prick capillary blood with plasma.

[f] 95%CI.

[g] Measurement of IP-10 LFA using plasma specimen.

[h] Including reagents, transport and staff.

[i] Cost per person-year included ART drugs (assuming complete linkage-to-care), imaging and laboratory tests (VL testing was excluded) and surveillance, and treatment of opportunistic infections including tuberculosis and cotrimoxazole prophylaxis.

[j] The model assumed that individuals who did not achieve VL suppression after receiving EAC switched to second-line ART.

[k] Cost of a new HIV infection in the first year also included the HIV testing and the counselling.

[l] The model assumed that the cost of a VF episode was for one year.

Abbreviations: AIDS: acquired immunodeficiency syndrome, ART: antiretroviral therapy, DALY: disability-adjusted live years, EAC: enhanced adherence counselling, HIV: human immunodeficiency virus, ICER: incremental cost-effectiveness ratio, LFA: lateral flow assay, VF: virological failure, VL: viral load.

Sources:

Relatório Anual das Actividades Relacionadas a HIV-SIDA 2021. Maputo, República de Moçambique: 2022.

Life expectancy at birth, total (years) - Mozambique | Data. Available at: https://data.worldbank.org/indicator/SP.DYN.LE00.IN?locations=MZ. Accessed 10 November 2022.

McCluskey SM, Boum Y, Musinguzi N, et al. Appraising Viral Load Thresholds and Adherence Support Recommendations in the World Health Organization Guidelines for Detection and Management of Virologic Failure. J Acquir Immune Defic Syndr 2017; 76:183. Available at: /pmc/articles/PMC5597473/. Accessed 10 November 2022.

Poorolajal J, Hooshmand E, Mahjub H, Esmailnasab N, Jenabi E. Survival rate of AIDS disease and mortality in HIV-infected patients: a meta-analysis. Public Health 2016; 139:3–12.

WHO methods and data sources for global burden of disease estimates 2000-2019. Geneva: 2020. Available at: http://www.who.int/gho/mortality_burden_disease/en/index.html. Accessed 10 November 2022.

Fajardo E, Metcalf CA, Chaillet P, et al. Prospective evaluation of diagnostic accuracy of dried blood spots from finger prick samples for determination of HIV-1 load with the NucliSENS Easy-Q HIV-1 version 2.0 assay in Malawi. J Clin Microbiol 2014; 52:1343–1351. Available at: https://journals.asm.org/doi/10.1128/JCM.03519-13. Accessed 10 November 2022.

Swannet S, Decroo T, de Castro SMTL, et al. Journey towards universal viral load monitoring in Maputo, Mozambique: many gaps, but encouraging signs. Int Health 2017; 9:206. Available at: /pmc/articles/PMC5881256/. Accessed 10 November 2022.

Diress G, Dagne S, Alemnew B, Adane S, Addisu A. Viral Load Suppression after Enhanced Adherence Counseling and Its Predictors among High Viral Load HIV Seropositive People in North Wollo Zone Public Hospitals, Northeast Ethiopia, 2019: Retrospective Cohort Study. AIDS Res Treat 2020; 2020. Available at: https://pubmed.ncbi.nlm.nih.gov/32373359/. Accessed 10 November 2022.

Bvochora T, Satyanarayana S, Takarinda KC, et al. Enhanced adherence counselling and viral load suppression in HIV seropositive patients with an initial high viral load in Harare, Zimbabwe: Operational issues. PLoS One 2019; 14. Available at: https://pubmed.ncbi.nlm.nih.gov/30721229/. Accessed 10 November 2022.

Korenromp EL, Gobet B, Fazito E, Lara J, Bollinger L, Stover J. Impact and Cost of the HIV/AIDS National Strategic Plan for Mozambique, 2015-2019—Projections with the Spectrum/Goals Model. PLoS One 2015; 10:e0142908. Available at: https://journals.plos.org/plosone/article?id=10.1371/journal.pone.0142908. Accessed 23 October 2022.

Pastor L, Casellas A, Carrillo J, et al. IP-10 Levels as an Accurate Screening Tool to Detect Acute HIV Infection in Resource-Limited Settings. Sci Rep 2017; 7. Available at: https://pubmed.ncbi.nlm.nih.gov/28808319/. Accessed 12 February 2022.
