## Supplementary Table 2 for "Field performance and cost-effectiveness of a point-of-care triage test for HIV virological failure in Southern Africa"

**Supplementary Table 2. Socio-demographic and clinical characteristics associated with virological failure (VF) (viral load >1,000 copies/mL) among study participants (n=209). Crude odds ratios from logistic regression analyses.**

| | N (%) or median (IQR) | | | | | | cOR | 95%CI | p-value |
|---|---|---|---|---|---|---|---|---|---|
| | Total (n=209) [a] | | VL≤1000 copies/mL (n=171) | | VL>1000 copies/mL (n=37) | | | | |
| **Biological sex** | | | | | | | | | |
| Male | 34 | (16.35) | 28 | (16.28) | 6 | (16.22) | Ref. | | |
| Female | 175 | (83.73) | 144 | (83.72) | 31 | (83.78) | 0.96 | 0.38-2.43 | 0.925 |
| **Age (years)** | 38 | (31-44) | 37 | (31-44) | 39 | (34-46) | 1.03 | 0.98-1.07 | 0.246 |
| **Employment** | | | | | | | | | |
| Employed | 43 | (20.57) | 38 | (22.09) | 5 | (13.51) | Ref. | | |
| Part-time | 21 | (10.10) | 14 | (8.14) | 7 | (18.92) | 3.62 | 1.03-12.70 | **0.045** [b] |
| Not employed | 145 | (69.38) | 120 | (69.77) | 25 | (67.57) | 1.48 | 0.55-3.99 | 0.437 |
| **Marital status** | | | | | | | | | |
| Married [c] | 58 | (27.75) | 51 | (29.65) | 7 | (18.92) | Ref. | | |
| Single | 139 | (66.51) | 112 | (65.12) | 27 | (72.97) | 1.68 | 0.70-4.02 | 0.244 |
| Divorced | 7 | (3.35) | 7 | (4.07) | 0 | (0.00) | 0.46 | 0.02-8.86 | 0.605 |
| Widowed | 5 | (2.39) | 2 | (1.16) | 3 | (8.11) | 9.61 | 1.60-57.85 | **0.013** [b] |
| **Educational level** | | | | | | | | | |
| No primary school | 10 | (4.78) | 6 | (3.49) | 4 | (10.81) | Ref. | | |
| Primary school | 135 | (64.59) | 112 | (65.12) | 23 | (62.16) | 0.30 | 0.08-1.09 | 0.067 [b] |
| Secondary school | 58 | (27.75) | 49 | (28.49) | 9 | (24.32) | 0.28 | 0.07-1.11 | 0.070 [b] |
| Post-secondary school | 2 | (0.96) | 1 | (0.58) | 1 | (2.70) | 1.44 | 0.11-18.73 | 0.779 |
| University | 4 | (1.91) | 4 | (2.33) | 0 | (0.00) | 0.16 | 0.01-3.78 | 0.256 |
| **BMI** [d] (n=202) | | | | | | | | | |
| Normal weight | 58 | (28.71) | 46 | (27.88) | 12 | (32.43) | Ref | | |
| Underweight | 2 | (0.99) | 1 | (0.61) | 1 | (2.70) | 3.72 | 0.35-38.92 | 0.273 |
| Overweight | 63 | (31.19) | 50 | (30.30) | 13 | (35.14) | 0.99 | 0.42-2.37 | 0.990 |
| Obesity | 79 | (39.11) | 68 | (41.21) | 11 | (29.73) | 0.62 | 0.26-1.51 | 0.296 |
| **Previous confirmed COVID-19 diagnosis** | 7 | (3.35) | 5 | (2.91) | 2 | (5.41) | 2.14 | 0.46-9.99 | 0.331 |

| | | | | | | | | | |
|---|---|---|---|---|---|---|---|---|---|
| **Previous COVID-19 hospitalization** | 1 | (0.48) | 1 | (0.58) | 0 | (0.00) | 1.52 | 0.06-38.16 | 0.797 |
| **Pregnancy** (n=175) | | | | | | | | | |
| No or unknown [e] | 170 | (97.14) | 141 | (97.92) | 29 | (93.55) | Ref. | | |
| Yes | 5 | (2.86) | 3 | (2.08) | 2 | (6.45) | 3.43 | 0.64-18.21 | 0.149 |
| **Hypertension** | 22 | (10.53) | 17 | (9.88) | 5 | (13.51) | 1.50 | 0.54-4.21 | 0.438 |
| **Diabetes** | 3 | (1.44) | 2 | (1.16) | 1 | (2.70) | 2.80 | 0.36-21.89 | 0.326 |
| **Hepatitis B** (n=65) | 3 | (4.62) | 1 | (2.44) | 2 | (8.33) | 3.00 | 0.37-24.23 | 0.303 |
| **Epilepsy** | 3 | (1.44) | 2 | (1.16) | 1 | (2.70) | 2.80 | 0.36-21.89 | 0.326 |
| **Symptoms at the study visit** [f] | 4 | (1.91) | 3 | (1.74) | 1 | (2.70) | 1.99 | 0.28-13.93 | 0.488 |
| **Current TB** | 1 | (0.48) | 0 | (0.00) | 1 | (2.70) | 14.18 | 0.57-355.02 | 0.107 |
| **Previous or current TB treatment** | 51 | (24.40) | 39 | (22.67) | 12 | (32.43) | 1.66 | 0.77-3.56 | 0.195 |
| **WHO stage at ART initiation** | | | | | | | | | |
| Stage 1 or 2 | 164 | (78.47) | 136 | (79.07) | 28 | (75.68) | Ref. | | |
| Stage 3 or 4 | 45 | (21.53) | 36 | (20.93) | 9 | (24.32) | 1.25 | 0.55-2.83 | 0.598 |
| **WHO stage at the study visit** | | | | | | | | | |
| Stage 1 or 2 | 159 | (76.08) | 132 | (76.74) | 27 | (72.97) | Ref. | | |
| Stage 3 or 4 | 50 | (23.92) | 40 | (23.26) | 10 | (27.03) | 1.25 | 0.57-2.76 | 0.583 |
| **IP-10 LFA reading** [g] (n=208) | 16.2 | (11.9-22.8) | 14.6 | (11.3-20.0) | 24 | (17.6-36.3) | 2.38 [h] | 1.66-3.40 | **<0.001** |
| **Days since last VL** | 7 | (4-14) | 8 | (5-14) | 6 | (2-12) | 0.97 | 0.93-1.02 | 0.233 |
| **Years since ART initiation** | 6.5 | (4.2-9.4) | 6.33 | (4.2-9.0) | 7.50 | (4.8-11.1) | 1.09 | 0.99-1.20 | 0.072 |
| **Years since HIV diagnosis** | 7 | (4-10) | 6 | (4-9) | 8 | (5-12) | 1.11 | 1.01-1.22 | **0.026** |
| **Missed ART** [i] | | | | | | | | | |
| None | 182 | (87.08) | 158 | (91.86) | 24 | (64.86) | Ref. | | |
| At least once dose a month | 27 | (12.92) | 14 | (8.14) | 13 | (35.14) | 6.02 | 2.56-14.16 | **<0.001** |
| **Current ART regimen** | | | | | | | | | |
| TDF+3TC+DTG | 174 | (83.25) | 150 | (87.21) | 24 | (64.86) | Ref. | | |
| TDF+FTC+EFV | 21 | (10.05) | 14 | (8.14) | 7 | (18.92) | 3.18 | 1.19-8.46 | **0.021** |
| Others | 14 | (6.70) | 8 | (4.65) | 6 | (16.22) | 4.70 | 1.55-14.21 | **0.006** |

Legend of table:

[a] For those variables with missing values, the total number of observations included is indicated next to the name of the variable.

[b] This category was not included in the multivariable analysis because of the small n and because it has not the potential to be a factor included in the ART monitoring algorithm.

[c] This category includes married, civil union or legal partnership.

[d] BMI ranges: normal weight: 18.5-24.9 kg/m2, underweight: <18.5 kg/m2, overweight: 25-29.9 kg/m2; obesity: ≥30 kg/m2.

[e] All of them were unknown, except one.

[f] After COVID-19 screening, symptoms assessed were fever, night sweats, myalgia, fatigue, headache, diarrhoea and skin rash

[g] IP-10 LFA values were log-transformed for a better adjustment of skewed data.

[h] For OR calculation, IP-10 reading values were categorized by increments of 10-units. Therefore, an increase of 10 units in the IP-10 reading value corresponds to a 2.38 OR of having VF.

[i] Self-reported ART adherence.

Abbreviations: 3TC: Lamivudine, ART: antiretroviral therapy, BMI: body mass index, CI: confidence interval, cOR: crude odds ratio, DTG: dolutegravir, FTC: emtricitabine, EFV: efavirenz, IQR: interquartile range, LFA: lateral flow assay, Ref: reference category, TB: tuberculosis, TDF: tenofovir, VL: viral load.
