## Supplementary Table 3 for "Field performance and cost-effectiveness of a point-of-care triage test for HIV virological failure in Southern Africa"

**Supplementary Table 3. One-way sensitivity analysis of the baseline value of the incremental cost-effectiveness ratio (ICER) (US$1,195/DALY averted) in a high transmission scenario (1:4).** ICERs were estimated comparing Strategy 2c against Strategy 2b. Value for each parameter is substituted one by one. DALY: disability-adjusted live years, EAC: enhanced adherence counselling, ICER: incremental cost-effectiveness ratio, LFA: lateral flow assay, VF: virological failure, VL: viral load.

| | Difference in costs (US$) | Difference in DALYs = DALYs averted | ICER (US$/DALY averted) |
|---|---|---|---|
| **VF prevalence 10%** | 310,034 | 39.8 | 7,788 |
| **VF prevalence 31%** | -237,142 | 125.4 | Strategy 2c dominates [a] |
| **VL coverage 48%** | -116,520 | 373.6 | Strategy 2c dominates [a] |
| **VL coverage 77%** | 100,643 | -32.6 | Strategy 2b dominates [a] |
| **VL test sensitivity 81.1%** | 84,591 | 39.2 | 2,157 |
| **VL test sensitivity 94.4%** | 51,386 | 28.1 | 1,826 |
| **VL test specificity 96.1%** | -35,833 | 121.8 | Strategy 2c dominates [a] |
| **VL test specificity 98.8%** | 119321 | -9.33 | Strategy 2b dominates [a] |
| **IP-10 LFA sensitivity 78.1%** | 340,576 | -225.3 | Strategy 2b dominates [a] |
| **IP-10 LFA sensitivity 98.3%** | -82,630 | 183.2 | Strategy 2c dominates [a] |
| **IP-10 LFA specificity 36.1%** | -141,993 | 183.9 | Strategy 2c dominates [a] |
| **IP-10 LFA specificity 51.4%** | 312,606 | -157.7 | Strategy 2b dominates [a] |
| **VL testing cost 24.78 US$** | 73,703 | 32.9 | 2,241 |
| **VL testing cost 54.12 US$** | 64,949 | 32.9 | 1,975 |
| VL suppression after EAC 31.3% | 10,922 | 14.5 | 755 |
| VL suppression after EAC 66.4% | 151,055 | 61.7 | 2,450 |

Legend of table:

[a] A dominant scenario occurs when the estimated costs are lower and the outcomes are improved.
